# Dopaminergic therapy selectively amplifies hallucination susceptibility in patients with Parkinson’s disease with cortico-striatal hyperconnectivity

**DOI:** 10.64898/2026.09.01.26361921

**Authors:** Fosco Bernasconi, Sara Stampacchia, Lucas Burget, Jevita Potheegadoo, Marie Maradan, Selim Habiby Alaoui, Sabina Catalano Chiuvé, Dimitri Van De Ville, Vanessa Fleury, Paul Krack, Olaf Blanke

## Abstract

Dopamine replacement therapy (DRT) alleviates motor symptoms in Parkinson’s disease (PD) but can trigger hallucinations in a subset of patients, yet the neural basis of this selective vulnerability is unknown. Hallucinations are among the most disabling non-motor symptoms of PD, linked to social isolation, dementia and institutionalization. Using a validated robotic paradigm to induce and quantify hallucinations in real-time, combined with resting-state fMRI in a crossover On/Off DRT design, we studied patients with PD with (PD-H) and without (PD-nH) hallucinations. DRT selectively amplified sensitivity to robot-induced hallucinations in patients with pre-existing hallucinatory phenotype (PD-H, but not PD-nH) and was accompanied by cortico-striatal and large-scale network hyperconnectivity. Rather than supporting a uniform hallucinogenic effect of dopamine in PD, these findings indicate that DRT interacts with an intrinsic neural vulnerability that varies in patients. Prospective studies will establish whether this pharmacological–behavioural signature identifies patients at risk before clinical hallucinations emerge.

## Introduction

Parkinson’s disease (PD) is characterized by the loss of dopaminergic neurons in the substantia nigra pars compacta, causing striatal dopamine deficiency and Parkinsonian motor symptoms (Kouli et al., 2018; Olanow & Tatton, 1999). Dopamine replacement therapy (DRT) is an effective symptomatic treatment to improve motor symptoms of PD (Goetz et al., 2005; Pahwa et al., 2006; Schapira, 2007). Yet, DRT has also been associated with well-documented complications. For instance, higher dosage eventually leads to severe motor complications, including motor dyskinesias (Gurevich & Gurevich, 2010). DRT has also been associated with important non-motor symptoms such as hallucinations and psychosis, which impose a substantial burden on patients, families, and caregivers (Ffytche et al., 2017), contributing to social isolation, earlier need for home nursing, and increased mortality; hallucinations are associated with an increased risk of cognitive decline and dementia (Bejr-Kasem et al., 2021; Bernasconi et al., 2023; Goetz & Stebbins, 1993).

The role of the DRT in inducing hallucinations was first recognized with the introduction of levodopa and later dopaminergic agonists. At that time, hallucinations in PD were commonly regarded as a drug-induced phenomenon (Barbeau, 1969; Poewe, 2008). Longitudinal studies confirmed that elevated levodopa-equivalent dosage was associated with increased risk of developing structured visual hallucinations (Forsaa et al., 2010; Zhu et al., 2013). Dopaminergic drugs have also been associated with minor hallucinations, which include presence and passage hallucinations as well as visual illusions (Fénelon et al., 2011; Kataoka & Ueno, 2015). Subsequent studies confirmed that early-stage PD patients have an increased risk of developing hallucinations with dopamine agonists (Morgante et al., 2012), and that these hallucinations often decrease in frequency/severity, or even disappear, after a dose reduction or an interruption of dopaminergic treatments (Friedman & Sienkiewicz, 1991; Mendis et al., 1996). Dopamine has also been associated with hallucinations in healthy individuals and psychiatric patients (Laruelle & Abi-Dargham, 1999; Reith et al., 1994), and with hallucination-like experiences in animals (Schmack et al., 2021).

However, other observations directly challenge this direct association between dopaminergic drugs and hallucinations. Historical data show that PD patients reported hallucinations well before the introduction of DRT (Gauthier et al., 1971; Rondot et al., 1984), and hallucinations can be observed today in PD patients who have not yet received any medication (Ala et al., 1997; Ballard et al., 1999; Pagonabarraga et al., 2016). Also, prior research has not found evidence of a simple dose–effect relationship between dopaminergic drugs and the development of hallucinations (Bernasconi et al., 2021, 2023; Fénelon et al., 2000; Sanchez-Ramos et al., 1996; Shergill et al., 1998). Accordingly, it has been argued that the occurrence of hallucinations reported in the early levodopa era may be related to the use of excessively high doses (Manford & Andermann, 1998; Pagonabarraga et al., 2024). Critically, Goetz and colleagues tested the relationship between intravenous dopamine administration and hallucinations, and found that intravenous levodopa injections and the consequent increase in blood levodopa levels did not induce any hallucinations in PD (Goetz et al., 1998), suggesting a more complex association between hallucinations and DRT.

Based on these data, the relationship between DRT and hallucinations and the involved neural mechanisms is still unclear and has been described as one of “the most controversial aspects” of this therapy (Ravina et al., 2007). One methodological aspect that may contribute to this controversial evidence might arise from the challenges associated with the investigation and quantification of hallucinations. The assessment of hallucinations predominantly relies on self-reports and their subsequent interpretation by researchers and clinicians, which carries well-documented shortcomings (Adler, 1973; Rosenthal & Fode, 1963). Verbal descriptions tend to capture hallucinations only partially, and are prone to both participant and experimenter biases, which may be particularly pronounced in clinical populations (Ravina et al., 2007). Moreover, hallucinations may have occurred at different time points before the interview (hours, days, weeks), rendering recall and related stratification difficult (Bernasconi et al., 2022; Rogers et al., 2021). All these considerations suggest that it is challenging to quantify whether DRT amplifies patients’ tendency to experience hallucinations. The clinical paradigm of choice for testing DRT effects is a within-subjects On-Off DRT design, in which medicated patients undergo dopamine withdrawal for approximately 12 hours (Langston et al., 1992). However, within such a short time window, the possibility to observe hallucinations and their changes in occurrence (e.g., frequency) is unlikely and might lead to false results. Therefore, the need for an experimental approach allowing to induce hallucinations and quantify changes in hallucination sensitivity within this short On/Off DRT window is critical.

To investigate the role of DRT on hallucinations and the associated neural mechanisms in PD, we used a novel patented method allowing to overcome the above mentioned difficulties in quantifying hallucinations (Bernasconi et al., 2022); the method demonstrated that a clinically relevant minor hallucination, presence hallucinations (PH), can be induced experimentally (i.e., robot-induced PH (riPH) or PH-like experiences) using repetitive somatomotor stimulation controlled by a robotic device. When applied in a conflicting somatomotor stimulation paradigm, this procedure has been shown to elicit riPH in patients with PD (Bernasconi et al., 2021; Potheegadoo, Duong Phan Thanh, et al., 2026) and in healthy participants (Blanke et al., 2014; Dhanis et al., 2026; Serino et al., 2021). Furthermore, it has been shown that PD patients with spontaneous hallucinations experienced in daily life (PD-H) are more sensitive to the riPH procedure than patients without hallucinations (PD-nH), whose sensitivity is comparable to that of healthy participants.

Here, we combined the riPH procedure and resting-state fMRI in a cross-over design to assess riPH sensitivity in the On and Off DRT states, in groups of non-demented PD patients with or without spontaneous hallucinations (PD-H; PD-nH) (Figure 1). Our data show that (1) patients with pre-existing hallucinatory traits (PD-H) and under their regular DRT intake (On state) show increased sensitivity to the riPH procedure compared with the Off state (no DRT medication intake). (2) This DRT-induced change in riPH sensitivity was not observed in patients without the hallucinatory trait (PD-nH). (3) Moreover, the increased sensitivity to riPH in patients with pre-existing hallucinatory trait (PD-H was associated with a pattern of cortico-striatal and large-scale network hyperconnectivity that (4) correlated with hallucinatory phenotype and sensitivity.

**Figure 1.**
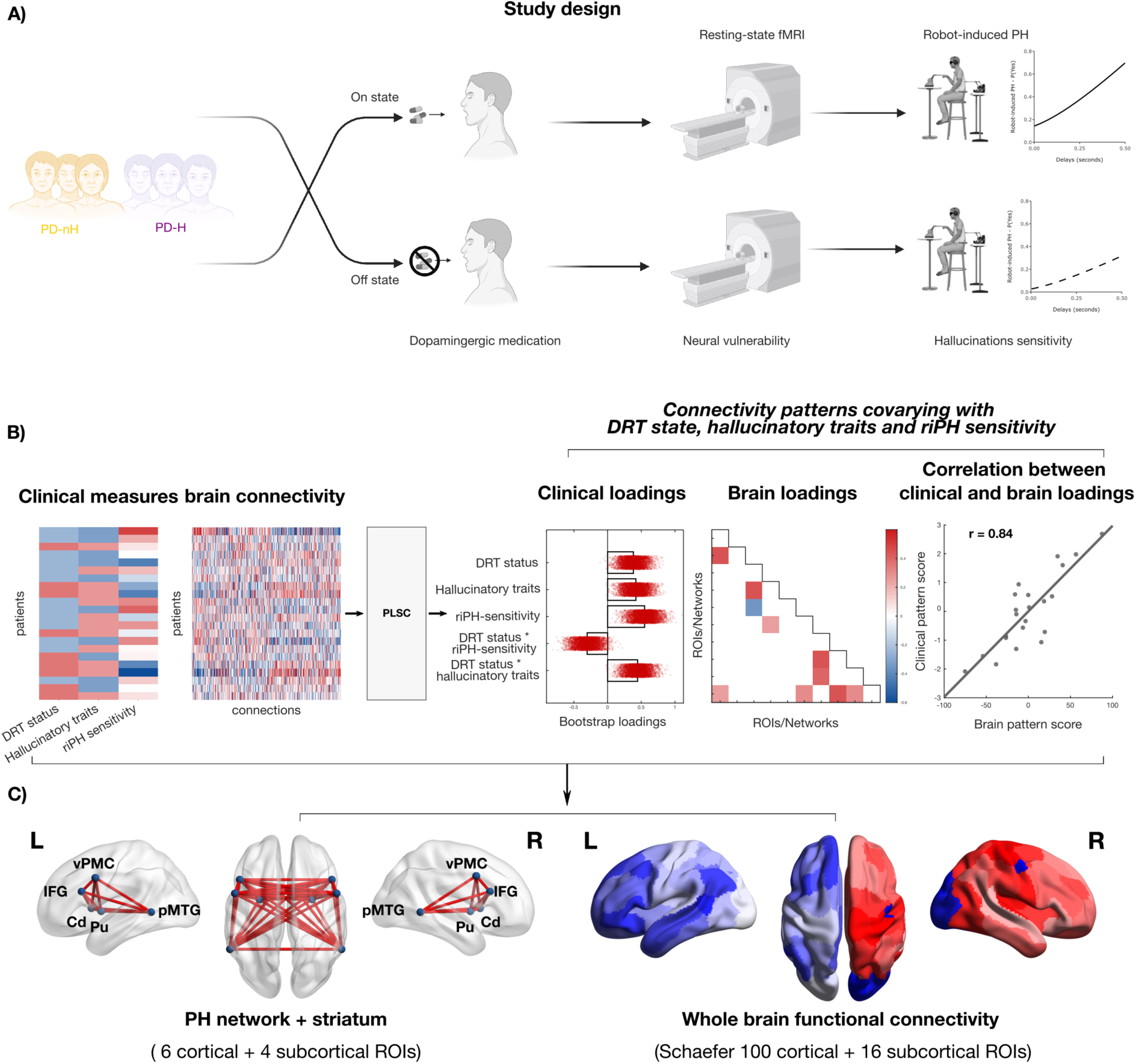
Mapping dopamine-induced hallucination sensitivity in Parkinson’s Disease. **A.** All patients with PD underwent the same crossover experimental design. In On state, participants took their regular DRT. In Off state, participants skipped the morning DRT. For both sessions, participants underwent a resting-state fMRI to assess the neural activity and a somatomotor robotic task to assess the riPH sensitivity. The order of the two sessions was pseudo-randomized across participants. **B.** Brain-behaviour associations were assessed using multivariate Partial Least Squares Correlations (PLSC) analyses to identify connectivity patterns covarying with DRT state, hallucinatory phenotype, and riPH sensitivity – that is, the connectivity patterns underlying dopamine-induced hallucination sensitivity. **C.** Two separate PLSC analyses were run: one restricted to regions previously associated with hallucination sensitivity (cortical and subcortical PH network (Bernasconi et al., 2021), and the striatum), and a second, data-driven, whole-brain analysis (Schaefer 100 cortical parcellation (Schaefer et al., 2018) plus 16 subcortical ROIs).

## Results

### Demographical, clinical, and neuropsychological data

Based on the semi-structured interview for the identification and characterization of psychotic phenomena in PD (see methods for details), we stratified the 20 patients with PD into those with hallucinations (PD-H; N = 11) and those without any hallucinations (PD-nH; N = 9) in daily life (i.e., hallucinatory trait: yes/no) (see supplementary materials for more details). Clinical-demographic data did not show any significant (all p-values > 0.05) differences in age, disease duration, or sex between the two subgroups of patients (Table 1). In line with previous work (Bernasconi et al., 2021, 2023; Potheegadoo, Duong Phan Thanh, et al., 2026; Stampacchia et al., 2026), no difference in the daily dosage of DRT, severity of motor impairment – assessed through the MDS-UPDRS Part 3 (Goetz et al., 2005), and cognitive functioning – assessed through the PD-CRS (Pagonabarraga et al., 2008) and MoCA (Nasreddine et al., 2005) was observed between the two patient sub-groups. In addition, there was no difference in the laterality of the onset of the motor symptoms (p-value = 1) nor in the overall onset laterality among all patients (left vs. right onset; p-value = 0.49).

**Table 1.** Clinical and demographic variables of the PD patients tested, divided by patients with hallucinations (PD-H), and patients without hallucinations (PD-nH). Values in the table represent the group’s mean, SD is indicated in brackets. LEDD-DA: Levodopa Equivalent Daily Dose - Dopamine Agonists; ; Global LEDD = Global daily Levodopa Equivalent Dose (all drugs combined). MoCA = Montreal Cognitive Assessment, assessing global cognitive function; MDS-UPDRS III = Movement Disorder Society–Unified Parkinson’s Disease Rating Scale, Part III, assessing motor symptom severity; H&Y = Hoehn and Yahr scale, assessing disease stage; PD-CRS = Parkinson’s Disease–Cognitive Rating Scale, assessing PD-specific cognitive impairment; Time Spent with Dyskinesias and impact with dyskinesias measured during the On DRT obtained from MDS-UPDRS Part 4.1. Neuropsychiatric fluctuations = presence of neuropsychiatric fluctuations (part III of the Ardouin Scale of Behavior in PD (Rieu et al., 2015).

|  | PD-H | PD-nH | p-values |
| --- | --- | --- | --- |
| Age (years) | 65.7 (9.0) | 60.8 (1.9) | 0.124 |
| Sex (M/F) | 8 / 3 | 7 / 2 | 1.000 |
| Disease duration (years) | 11.8 (3.0) | 11.8 (3.8) | 0.979 |
| LEDD-DA (mg/day) | 166.8 (150.0) | 171.9 (140.5) | 0.939 |
| Global LEDD (mg/day) | 1120.3 (493.1) | 1021.3 (203.4) | 0.581 |
| MDS-UPDRS III On | 23.5 (11.4) | 22.8 (10.2) | 0.877 |
| MDS-UPDRS III Off | 43.5 (6.9) | 41.3 (17.6) | 0.717 |
| H&Y On | 2.05 (0.151) | 1.94 (0.300) | 0.356 |
| H&Y Off | 2.2 (0.422) | 2.22 (0.363) | 1.00 |
| MoCA On | 27.6 (1.6) | 27.9 (1.4) | 0.709 |
| Total PD-CRS On | 96.5 (12.9) | 102.2 (11.2) | 0.287 |
| Total PD-CRS Off | 99.2 (10.1) | 102.2 (13.0) | 0.403 |
| Time dyskinesia | 1.0 (1.3) | 0.8 (1.2) | 0.694 |
| Impact dyskinesia | 0.636 (1.21) | 1 (1.273) | 0.47 |
| Neuropsychiatric fluctuations | 5.27 (3.52) | 6.7 (3.8) | 0.36 |

### DRT improves motor functions

As expected, when comparing motor symptoms in the Off vs. On DRT state, we observed a significant improvement in the On state, as reflected by significantly lower scores on the MDS-UPDRS (Part 3) (main effect of medication: p-value < 0.001; interaction group x medication: p-value = 0.381) and no significant differences for the Hoehn & Yahr scale (main effect of medication; p-value = 0.085; interaction group x medication: p-value = 0.719). Cognitive functions (PD-CRS) were comparable and not significantly affected by the medication state (On vs. Off; main effect of medication; p-value = 0.559; interaction group x medication: p-value = 0.518).

### On DRT state increases the riPH sensitivity in PD-H, not in PD-nH

Previous work in PD showed that riPH depends on the degree of somatomotor conflict (i.e., the mismatch between the motor and proprioceptive-tactile cues during hand-poking movement performed with the front robot and the tactile feedback delivered by the back robot), with stronger somatomotor conflict leading to a higher probability of experiencing riPH (Bernasconi et al., 2021, see methods). In the present study, in each trial, participants performed somatomotor stimulation consisting of a total of 10 individual pokes while being exposed to a randomly chosen somatomotor delay (0, 0.25, or 0.5 seconds). After each trial, participants indicated whether they experienced riPH (Yes/No). Participants performed the task both in the On and Off DRT state (with the order randomly attributed across participants), on different days (Figure 1).

We investigated the effects of DRT state (On, Off), hallucinatory phenotype (PD-H, PD-nH), and somatomotor delay (0, 0.25, 0.5 seconds) on riPH (Figure 2A) and any interaction between the three terms (see Tables S1). Confirming previous findings (Bernasconi et al., 2021; Potheegadoo, Thanh, et al., 2026), our results show that the robotic procedure elicited riPH independently from the group and that it was positively modulated by the somatomotor delay (main effect of delay: p-value < 0.001). Critically, the present results show that the occurrence of riPH was modulated by DRT state, delay, and hallucinatory phenotype, as indexed by a significant interaction between the three terms (p-value = 0.044). In the post-hoc analyses (to disentangle the triple interaction), we investigated the effects of delay and DRT separately for PD-H and PD-nH. Our results show that while in PD-H the riPH sensitivity was modulated by the DRT state (main effect of DRT: p-value = 0.008; Figure 2B; see supplementary Tables S2), this was not the case for PD-nH (no main effect of DRT state: p-value = 0.918; Figure 2C; see supplementary Tables S3). These results show that the riPH procedure allows to identify two distinct response patterns to DRT. In PD-H, DRT increases the sensitivity to riPH, while it is not affected in PD-nH. For both groups, the increase of the somatomotor conflict resulted in a higher probability to experience riPH (PD-H, main effect of delay, p-value < 0.001; PD-nH, main effect of delay, p-value = 0.002). As a control analysis, we compared the riPH in the Off DRT state between PD-H and PD-nH; results showed no statistical difference between the two groups (p-value = 0.346; Figure 2D), suggesting that the two groups have a similar riPH sensitivity in the absence of DRT and showing that the DRT impact on riPH sensitivity is not due to baseline (Off DRT) differences.

**Figure 2.**
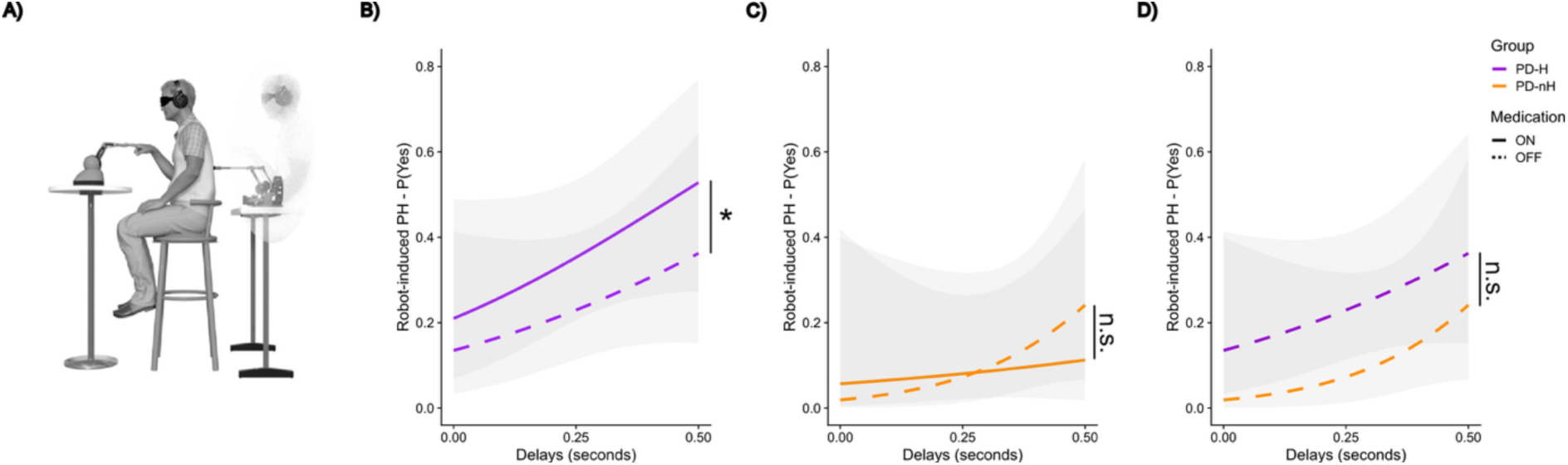
Dopaminergic medication in PD patients with hallucinatory trait is associated with higher sensitivity to riPH. **A.** Illustration of the somatomotor setup and procedure leading to riPH. **B.** Visualization of the riPH as a function of delay, On DRT state (continuous line) compared to the Off DRT state (dashed line) in PD-H. Asterisk indicates significant effect (p-value = 0.008). **C.** The model-derived predicted probabilities of riPH failed to show a significant difference in riPH, with an increased sensitivity in the On DRT state compared to the Off state, in PD-nH. **D.** In the Off DRT state the riPH did not differ significantly between PD-H and PD-nH (all p-values > 0.05). For subfigures B-D, the thicker line indicates the mean, and the error bars indicate 95% confidence intervals. n.s. indicate not significant.

Collectively, these data show that: i) robotic somatomotor conflict elicited riPH across all patients and showed a similar main effect of delay, ii) in PD-H patients, DRT boosted the riPH sensitivity in the On-state compared to the Off-state, while iii) PD-nH did not show a significant difference in riPH sensitivity between On-and Off-state; and iv) these two distinct response patterns to DRT are not due to baseline (Off DRT) differences. Providing experimental and quantitative evidence about the role of dopamine in hallucinations in PD, these robotic data show that the riPH procedure detects a heightened vulnerability to experience hallucinations after intake of a single habitual DRT dose in patients who report hallucinations in daily life (PD-H patients). That is, the same dopaminergic intervention differentially alters hallucination sensitivity depending on a patient’s pre-existing hallucinatory trait, suggesting a different underlying neural vulnerability between the two groups.

### Cortico-striatal hyperconnectivity underlies dopamine-enhanced sensitivity to riPH in PD-H

To determine the neural substrates of dopamine-modulated riPH sensitivity as a marker of hallucination vulnerability in PD, we conducted multivariate analyses (Partial Least Squared correlations (PLSC)) (Krishnan et al., 2011, 2011), extracting the patterns capturing shared variance (i.e. latent components) between functional connectivity and DRT status, hallucinatory phenotype and riPH sensitivity. We conducted two separate PLSC: these allowed us to investigate which functional connections (1) within a previously defined cortical PH-network (Bernasconi et al., 2021) and its connections to striatal regions (i.e., cortical-subcortical PH-network) and (2) across the whole-brain, are associated with DRT-driven (On>Off) enhanced hallucination sensitivity (delay-dependent riPH) in patients with and without hallucinatory trait (PD-H>PD-nH).

Concerning connectivity in the cortical-subcortical PH-network (inferior frontal gyrus (IFG), posterior middle temporal gyrus (pMTG), ventral pre-motor cortex (vPMC), putamen, caudate; all bilateral) the first PLSC analysis identified one significant latent component (LC; p-value=0.032) explaining 33.7% of the covariance between functional connectivity patterns and the combined behavioral and clinical measures. Behaviorally, this LC was characterized by a significant interaction of DRT status*riPH sensitivity, as well as DRT status*hallucinatory phenotype (PD-H>PD-nH), both carrying positive loadings (Fig. 3A). At the neural level, the LC reflected increased connectivity within the striatum (both between the caudate and the putamen and interhemispherically, i.e., left–right), which is part of the main dopaminergic pathway (Obeso et al., 2008), as well as increased PH-network – striatum connectivity (specifically R-caudate – R-IFG and R-caudate – L-vPMC), increased inter-hemispheric cortical connectivity within the PH-network (specifically R-pMTG – L-pMTG and R-vPMC – L-vPMC), and reduced connectivity between L-IFG and L-pMTG (Fig. 3B and Fig. 3C). Of the possible 45 connections tested, 12 reached significance (p-values < 0.05), with 11 reflecting hyperconnectivity, and only one hypoconnectivity (L-IFG – L-pMTG), indicating a pattern of predominant hyperconnectivity. Moreover, brain and behaviour scores were strongly correlated (r = 0.62), with the strongest hyperconnectivity observed in PD-H patients while On DRT (Fig. 3D). These results indicate that this LC captured a modulation of hyperconnectivity in the PH-network and dorsal striatum, driven primarily by heightened delay-dependent riPH sensitivity and pre-existing hallucinatory phenotype in the On vs. Off DRT state.

**Figure 3.**
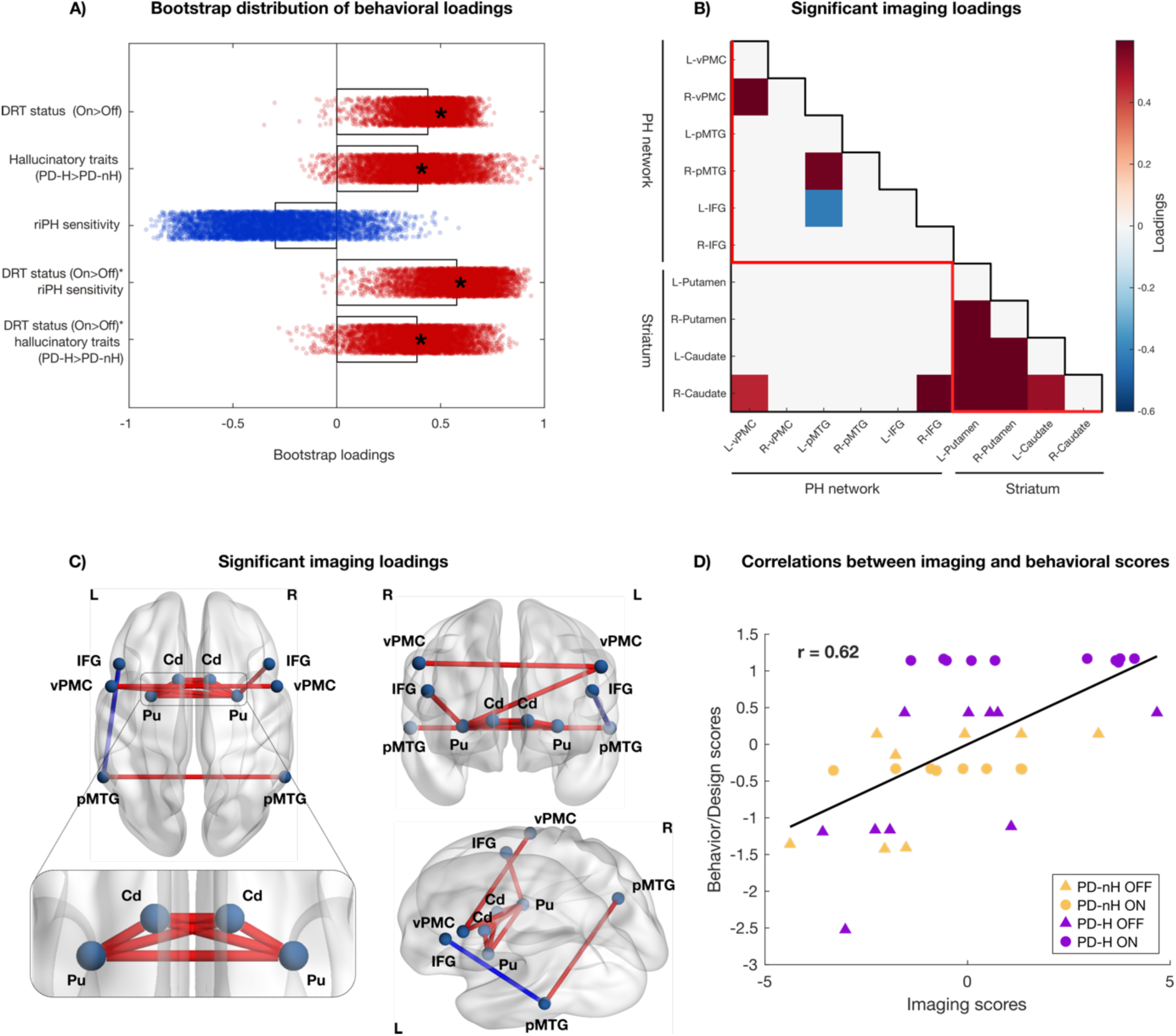
Connectivity signatures of dopaminergic medication and hallucination sensitivity in Parkinson’s disease: cortical and subcortical presence hallucination network. **A.** Bootstrap distribution of the behavioral loadings for the -significant latent component (LC; p = 0.032), explaining 33.7% of the covariance between functional connectivity of the subcortical and cortical PH-network, and the combined behavioral and clinical measures. Asterisks (*) indicate statistically significant loadings based on the 95% bootstrap confidence interval. red = positive loading; blue = negative loading. **B.** Matrix showing the significant imaging loadings (i.e., connections with statistically reliable loadings across bootstrap samples based on the 95% confidence interval), highlighting especially the striatal hyperconnectivity (lower right corner), as well as cortico-cortical connections (upper left corner). red = positive loading; blue = negative loading. IFG = inferior frontal gyrus; pMTG = posterior middle temporal gyrus; vPMC = ventral premotor cortex; L = left; R = right. **C.** Brain renders depicting the significant connections identified in the loading matrix (panel B), providing a cortical visualization of the network topography *in (B)*. Nodal strength was computed as the sum of edge weights for each ROI. Inset shows a zoom on subcortical connections. **D.** Correlation between imaging and behavioral (DRT state, hallucinatory phenotype, and riPH sensitivity) scores (r = 0.62).

### Hyperconnectivity in the somatomotor and default mode network underlies dopamine-modulated increased riPH sensitivity in PD-H

To further substantiate these fMRI findings, we performed a second PLSC analysis examining whole-brain functional connectivity without restricting the model to predefined regions, thereby enabling a fully data-driven analysis across the 100 ROIs of the Schaefer parcellation and 16 subcortical and striatal regions. This analysis identified a significant latent component (LC; p-value=0.040), accounting for 34.3% of the cross-covariance between functional connectivity and the combined behavioral and clinical measures. Consistent with the previous cortical-subcortical PH-network analysis, behaviourally this LC was characterized by a significant interaction of DRT state*riPH sensitivity and DRT state*hallucinatory phenotype (PD-H>PD-nH) (Fig. 4A). At the neural level, it was characterised predominantly by hyperconnectivity within the DMN and somatomotor (SMT) networks, as well as the following four between-network connectivities involving SMT and DMN: SMT-salience (SA), SMT-limbic (L), SA-L, DMN-dorsal attention (DA), and DMN-SA (Fig. 4B). Connectivity within the striatum almost reached significance (z = 1.414). Network-pair enrichment was assessed using a permutation-based test (5,000 permutations of network labels preserving network size), yielding a one-tailed z-score per network pair to correct for unequal network size (z > 1.645, p<.05, one-tailed). Most significant edges showed positive loadings (302/327, 92.4%); the smaller subset of negative loadings (25/327, 7.6%) was too sparse for reliable network-pair enrichment testing and is reported descriptively, alongside raw edge counts per network pair (Supplementary Fig. S1). Of note, regions with the highest imaging loadings overlapped with PH-network areas – including the left IFG, pMTG, and vPMC (Fig. 4C). Again, brain and behaviour scores were robustly correlated (r = 0.92), with the strongest brain loadings observed in patients with hallucinatory trait (PD-H) during the On DRT state (Fig. 4D). These fMRI results link a full brain LC – characterized predominantly by hyperconnectivity within DMN and somatomotor networks, and by increased connectivity between these networks and the DA, SA and L networks – to dopamine-enhanced hallucination sensitivity in PD patients with hallucinatory trait (PD-H). Overall, these fMRI results extend previous findings of cortico-striatal hyperconnectivity associated with dopamine-enhanced hallucination sensitivity (measured with riPH) in PD-H patients, showing that this cortical hyperconnectivity primarily involved DMN, SMT, and SA, consistent with a neural marker of dopamine-modulated hallucination vulnerability.

**Figure 4.**
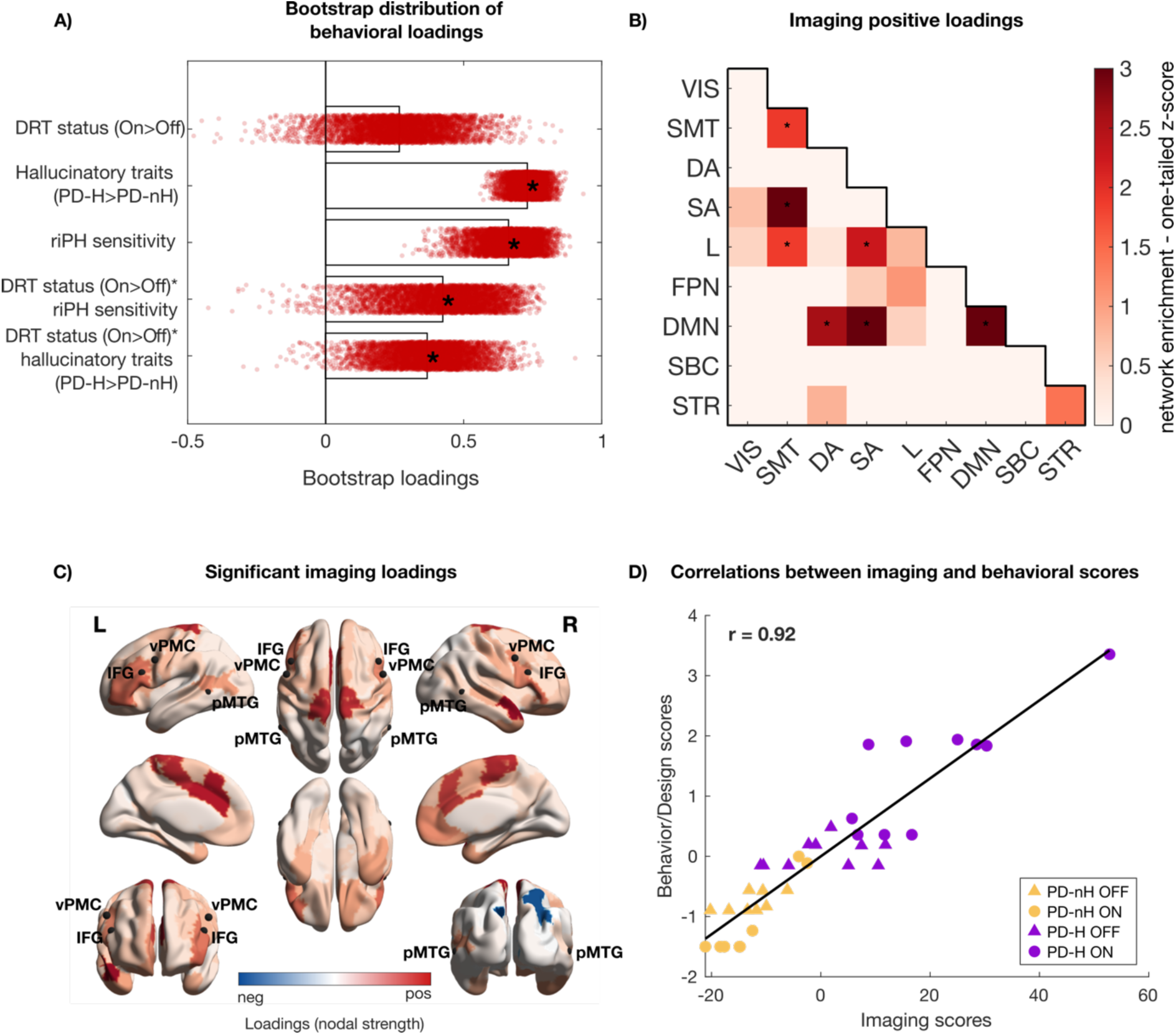
Connectivity signatures of dopaminergic medication and hallucination sensitivity in Parkinson’s disease: whole-brain analyses. **A.** Bootstrap distribution of the behavioral loadings for the first – and only-significant – latent component (LC; p = 0.040), which explained 34.3% of the covariance between whole-brain functional connectivity and the combined behavioral and clinical measures. Asterisks (*) indicate statistically significant loadings based on the 95% bootstrap confidence interval. Red = positive loading; blue = negative loading. **B.** Matrix shows the network-pair enrichment of positive significant imaging loadings. * indicate network pairs with significant enrichment above chance (z > 1.645, p<.05, one-tailed). Matrix is organized by the *7* canonical resting-state networks (RSNs), subcortical regions, and striatum: VIS = Visual Network; SMT = Somatomotor Network; DA = Dorsal Attention Network; SA = Salience Network; L = Limbic Network; FPN = Fronto-Parietal Network; DMN = Default Mode Network; SBC = Subcortical regions (including left and right thalamus, hippocampus, globus pallidum, amygdala and pons); STR = Striatum (including left and right putamen, caudate and nucleus accumbens). **C.** Brain renders showing the nodal strength of the significant loading matrix, providing a cortical visualization of the network topography in B. For visualization purposes, we overlaid the center of mass of the PH-network ROIs used in previous analyses. The regions with the highest nodal strength partially overlapped with the PH-network, particularly in the bilateral IFG, pMTG, and vPMC, with a predominance in the left hemisphere. IFG = inferior frontal gyrus; pMTG = posterior middle temporal gyrus; vPMC = ventral premotor cortex. D) Correlation between imaging and behavioral (DRT state, hallucinatory phenotype and robot sensitivity) scores (r = 0.92).

## Discussion

While DRT alleviates motor symptoms in PD, the mechanisms underlying its association with hallucinations, one of the most disabling non-motor symptoms in PD, remains poorly understood. In the present study, we assessed hallucination sensitivity using a novel and validated hallucination-induction procedure (riPH) and collected resting-state fMRI in the On and Off DRT states in PD-H and PD-nH patients, to determine whether patients depending on their pre-existing hallucinatory phenotype are differently susceptible to DRT-facilitated hallucinations and to identify any underlying neural vulnerability. First, we provide evidence that PD-H are more sensitive to the riPH procedure in On vs. Off DRT (Figure 2). In contrast, in PD-nH patients, the riPH sensitivity was not modulated by DRT, showing a different degree of hallucination vulnerability between the two groups. Second, we demonstrate that increased riPH sensitivity observed in PD-H patients in the On DRT state is driven by within-striatal, cortico-striatal and cortico-cortical hyperconnectivity (Figure 3). Third, whole-brain analysis corroborated and extended these findings by revealing increased intra-and inter-network connectivity among the DMN, SMT and SA networks, again depending on DRT-state, pre-existing hallucinatory phenotype and riPH sensitivity (Figure 4). Collectively, these results show that cortical and subcortical hyperconnectivity is a potential neural vulnerability signature for dopamine-induced hallucinations in PD, uncovered by the riPH procedure.

While early clinical observations have linked DRT to hallucinations in PD (Barbeau, 1969; Poewe, 2008), through dose-dependent associations (Fénelon et al., 2011; Forsaa et al., 2010; Friedman & Sienkiewicz, 1991; Kataoka & Ueno, 2015; Mendis et al., 1996; Morgante et al., 2012; Zhu et al., 2013), in healthy, psychiatric, and animal models (Cassidy et al., 2018; Laruelle & Abi-Dargham, 1999; Reith et al., 1994; Schmack et al., 2021), other evidence challenges a direct causal role: hallucinations have been reported in unmedicated PD patients (Ala et al., 1997; Ballard et al., 1999; Gauthier et al., 1971; Pagonabarraga et al., 2014; Rondot et al., 1984), a clear dose–effect relationship has not been consistently found (Bernasconi et al., 2021, 2023; Manford & Andermann, 1998; Pagonabarraga et al., 2024; Potheegadoo, Duong Phan Thanh, et al., 2026; Sanchez-Ramos et al., 1996; Shergill et al., 1998; Stampacchia et al., 2026), and, critically, experimental intravenous dopamine administration failed to induce hallucinations(Goetz et al., 1998). The present results show that although having received a similar dose of dopaminergic treatment, DRT increases the sensitivity to hallucinations in one phenotype of patients (PD-H) but has no effect in another (PD-nH), supporting a model in which dopamine interacts with a pre-existing vulnerability for hallucinatory experiences rather than supporting a uniform hallucinogenic effect of dopamine, reconciling conflicting earlier findings in the literature.

Our findings were made possible by the experimental induction and real-time quantification of riPH sensitivity using somatomotor conflict controlled through the robotic procedure. This allowed us to experimentally determine each patient’s sensitivity in the riPH procedure within the short Off (washout) period (∼12 hours). This was not possible in previous studies. While the administration of DRT has been linked to hallucinations in PD, based on free reports (Fénelon et al., 2011; Forsaa et al., 2010; Kataoka & Ueno, 2015; Morgante et al., 2012; Zhu et al., 2013), previous studies attempting to capture whether the administration of DRT triggered hallucinations within a short and controlled timed frame (after intravenous administration) did not succeed (Goetz et al., 1998). We argue that this was possibly the case because former authors lacked methods to quantify hallucination sensitivity (in real-time) independently of spontaneous symptom occurrence or free patient report (for discussion see (Bernasconi et al., 2022)). The present experimental approach to hallucinations not only allows to show that DRT increases the sensitivity to hallucinations, but it also shows that this effect is specific for patients (PD-H) with a pre-existing hallucinatory phenotype, suggesting that differences in neural vulnerability may have increased their sensitivity to DRT and hallucinations.

Neurally, our results show that the increased sensitivity to riPH observed in PD-H during the On DRT state is associated with two neural signatures: cortico-striatal hyperconnectivity and large-scale network hyperconnectivity. Striatal dopamine has been associated with hallucination-like experiences (Cassidy et al., 2018; Schmack et al., 2021). Based on the present data, we speculate that the heightened striatal connectivity may reflect a dopaminergic overstimulation mechanism (Lee et al., 1978), which was previously linked to hallucinations in PD (de Celis Alonso et al., 2015; Haslinger et al., 2001). Thus, this hyperstimulation of the striatum induced by the DRT could lead to hallucinations in vulnerable patients (Dave et al., 2019; Dujardin & Sgambato, 2020; Jaakkola et al., 2017; Kiferle et al., 2014; Lenka et al., 2015; Papapetropoulos, 2006; Ravina et al., 2012; Zahodne & Fernandez, 2008). Our neural results also show that the dopamine-enhanced hallucination sensitivity in PD-H was further associated with increased functional connectivity beyond the striatum: between the striatum and the PH-network. Prior work from our group has shown that increased connectivity in the PH-network predicts PD-H phenotype (Bernasconi et al., 2021). The results of the current work extend these findings by showing that cortico-striatal connectivity is upregulated by dopaminergic medication. Our results also showed a large-scale network hyperconnectivity, including the DMN and SMT (both overlapping with the PH-network) and between both these networks and the SA network. Altered connectivity within and between the DMN and SA has been implicated in hallucinations in PD (Shine et al., 2011, 2014), and increased dopamine levels have been shown to increase the coupling between the DMN and SA (Delaveau et al., 2010; Zhong et al., 2019). While we corroborate the role of DMN and SA in PD hallucinations in general, our results provide new evidence for a link between these cortical alterations and dopaminergic susceptibility to hallucinations in PD. Our data further extend this neural vulnerability to SMT. Interestingly, the SMT has been primarily linked to the core motor pathophysiology of PD and is a target of DRT (Tessitore et al., 2014), our SMT findings, applying a somatomotor induction method, extend previous work on hallucinations in PD, having shown that somatomotor (Bernasconi et al., 2021), but also cerebello-cortical motor systems (Stampacchia et al., 2026) play a critical role in hallucinations. We here extend these findings by providing a new link between the SMT network and DRT-related hallucinations in PD. Finally, previous evidence has highlighted that during visual misperceptions, primary visual regions become hyperconnected with the DMN (Shine et al., 2015). Here, we extend those findings by showing that riPH is also associated with hyperconnectivity between the DMN and sensory cortex, but with SMT instead of visual areas. Critically, a similar dose of DRT does not lead to either of these hyperconnectivity patterns in the present control group of PD patients (PD-nH), possibly because of a reduced cortico-striatal vulnerability in the described networks.

In conclusion, our study provides quantitative evidence concerning the “most controversial aspect” (Ravina et al., 2007) of DRT. We demonstrate that DRT can increase hallucination sensitivity in PD patients, but only in those having a pre-existing hallucinatory phenotype and neural vulnerability characterized cortico-striatal hyperconnectivity and large-scale network hyperconnectivity. This suggests that dopamine has not a uniform hallucinogenic effect in all patients with PD, but only in patients with the described neural vulnerability that can be detected based on the riPH procedure. From a clinical perspective, these findings may help shift the interpretation of hallucinations from a simple dose-dependent adverse effect of DRT toward an interaction between treatment and individual neural vulnerability. Future longitudinal studies should investigate whether DRT-enhanced riPH sensitivity and the related cortico-striatal and large-scale network hyperconnectivity pattern are a potential biomarker to identify patients at risk of dopamine-induced hallucinations, opening avenues to pre-emptive stratification of treatment side-effect risk before psychotic symptoms emerge.

### Limitations

Our findings should be considered in the context of the following limitations. First, the overnight withdrawal used to define the Off DRT state does not fully eliminate the long-duration response to chronic levodopa treatment (Cilia et al., 2020; Nutt & Holford, 1996). However, we only included patients experiencing motor and neuropsychiatric fluctuations, which is important when interpreting the On vs Off DRT contrast, as these fluctuations are associated with an increasing contribution of the short-duration response variations to levodopa, while the long-duration response may become relatively less effective in smoothing these acute variations (Nutt & Holford, 1996). Thus, although the Off condition cannot be considered fully treatment-free because a residual long-duration response is likely to persist, the selection of fluctuating patients makes it likely that the experimental On vs Off DRT manipulation captured a clinically meaningful short-duration dopaminergic effect. Second, the sample size (N = 20) may appear modest, yet it should be considered in light of the demanding nature of the pharmacological within-subject protocol: each patient completed extensive testing across both On and Off DRT states, including supervised medication withdrawal and repeated experimental sessions. Testing 20 patients under these constraints already represents a substantial undertaking, and future studies will be valuable for replicating and extending these findings in larger cohorts. Third, future studies could examine different levels of On-DRT states (e.g., 50%, 100%, and 150% of the prescribed dopaminergic drug dose) to examine dose-dependent changes, which could be captured by the riPH procedure. Fourth, longitudinal studies could further extend these findings by examining whether sensitivity to PH-like experience in the de novo patients (prior to DRT) predicts the subsequent development of hallucinations over time. Fifth, future studies might use implicit behavioral measures to quantify hallucinations (Albert et al., 2024) and also use MEG/EEG to assess whether DRT alters oscillatory activity that have been associated with hallucinations in PD (Bernasconi et al., 2025). Finally, it should be noted that although our results show that dopamine has an important role in hallucinations in PD, it is unlikely to be the only neurotransmitter involved in the generation of hallucinatory perceptions. Hence, growing evidence shows that hallucinations might be related to multiple neurotransmitters such as serotonin and acetylcholine (Collerton et al., 2023; Ffytche et al., 2017; Pagonabarraga et al., 2024; Stampacchia et al., 2026). For instance, hallucinations in PD may arise from a serotonin-dopamine imbalance (Stahl, 2016). Future studies should address this open question by studying the effects of different neurotransmitter systems and their interactions on the genesis of hallucinations.

## Materials and Methods

### Participants

Twenty-two individuals participated in this study. Individuals were selectively recruited during routine outpatient consultations at the Movement Disorders Clinic of InselSpital Bern (Switzerland) and the Geneva University Hospital (HUG, Switzerland) by movement disorders specialists. Inclusion criteria were age above 18 years, PD diagnosis (Hughes et al. 1992), presence of neuropsychiatric fluctuations (sum ≥3 on part III of the Ardouin Scale of Behavior in PD (Rieu et al. 2015) and ongoing DRT (Jost et al., 2023; Tomlinson et al., 2010). One patient was not able to perform any task in the Off state, and one patient had to be excluded because of excessive movement in the MRI. Each individual was interviewed regarding disease onset, medication history, current medications, and dosage (levodopa equivalent daily dose (LEDD) and levodopa-equivalent dose (LED) of dopaminergic agonists) (Jost et al., 2023; Tomlinson et al., 2010)(Table 1). Motor symptoms severity was assessed using the Movement Disorder Society – Unified Parkinson’s Disease Rating Scale part III scale (MDS-UPDRS-III) (Goetz et al., 2008), and disease severity and functional impact using the Hoehn & Yahr scale (H&Y) (Goetz et al., 2004; Hoehn & Yahr, 1967) during both the On medication and Off medication states. Also, cognition was examined during both On and Off state by an expert clinical neuropsychologist using the PD-specific cognitive functioning using Parkinson’s Disease – Cognitive Rating Scale (PD-CRS) (Pagonabarraga et al., 2008) – further details are reported in the ‘*Hallucinations and cognitive function assessment*’ paragraph. Global cognition was assessed using the Montreal Cognitive Assessment (MoCA) (Nasreddine et al., 2005). Individuals were divided into two sub-groups: patients with spontaneous hallucinations - PD- H (i.e., presence, passage hallucinations, visual illusions and/or pareidolias, auditory, tactile, or structured visual hallucinations (n = 11); see below for more details) and patients without any hallucinations PD-nH (n = 9). The two subgroups of patients were comparable in sex, disease duration, LEDD, and LED of dopaminergic agonists, motor symptoms severity (MDS-UPDRS-III), and cognition (MoCA, PD-CRS).

Exclusion criteria included a history of major psychiatric disorders, cerebrovascular disease, conditions known to impair mental status other than PD, and the presence of factors that prevented magnetic resonance imaging (MRI) scanning (for example, claustrophobia, MRI incompatible prosthesis). Patients with anatomical abnormalities in MRI or non-compensated systemic diseases (that is, diabetes and hypertension) were also excluded. All participants were on stable doses of dopaminergic drugs for at least 4 weeks before inclusion. No participant had used or was using antipsychotic medication. All subjects had normal or corrected-to-normal vision. Informed consent to participate in the study was obtained from all participants. The study was approved by the local cantonal ethics committee (Bern: #2021-01608; Geneva: # 2017-01852).

### Hallucinations and cognitive functions assessments

The presence and type of hallucinations experienced spontaneously by the participants were assessed by clinical neuropsychologists using a short questionnaire designed to assess the occurrence of hallucinations in patients with PD. Hallucination occurrence and frequency were assessed over the four weeks before inclusion in the study. The following hallucinations were evaluated for each participant: passage hallucinations (i.e. the feeling that someone, something, or a shadow passing or moving in the peripheral visual field), presence hallucinations (i.e. the feeling that someone is behind or close to you when no one is actually there), visual illusions/pareidolia (i.e. misperceptions in which a real visual stimulus is present but misinterpreted, such as perceiving figures or faces in ambiguous objects), complex visual hallucinations (VH) (i.e. vividly seeing objects, people, animals or scenes that are not present), auditory (i.e. clearly hearing sounds, voices, music, or noises in the absence of an external source) and tactile hallucinations (i.e. vivid sensations of touch, itching, burning, heat, cold, or abnormal skin sensations in the absence of physical stimulation). In total, seven patients reported passage hallucinations, five reported presence hallucinations, 6 reported visual illusions, 2 complex visual, 5 auditory and 3 tactile hallucinations. Cognition was assessed using the PD-CRS, a cognitive scale specifically designed to capture the whole spectrum of cognitive functions impaired over the course of PD. This battery comprises nine tasks explicitly designed for a brief and separate scoring procedure, including frontal–subcortical tasks (sustained attention, working memory, alternating and action verbal fluency, clock drawing, immediate and delayed free recall verbal memory) as well as posterior cortical tasks (confrontation naming, clock copying). The frontal– subcortical assessment score ranges from 0 to 114 points, the posterior assessment score from 0 to 20 points. The sum of the two scores is added to give the total score of the PD-CRS (0–134). Lower scores indicate lower cognitive functions (Table 1). In addition, global cognition was assessed using MoCA, only during the On-state and it was used to exclude dementia (total score < 22).

### Study design

Objectives of the study were to (i) investigate the role of dopaminergic medications (On vs. Off medication) on the induction of experimentally induced PH; (ii) assess potential differences in sensitivity to riPH in PD patients with and without hallucinations (*Group*: PD-H vs PD-nH); (iii) assess the role of somatomotor stimulation on riPH (; (iv) identify the neural networks associated with changes in sensitivity to riPH depending on the medication state. With this aim, PD patients were asked to perform our sensorimotor robotic task (see below for more details), in On dopaminergic medication and in Off dopaminergic medication. The two sessions were conducted in a random order across participants and on separate days (the order On/Off was not significantly different between PD-H and PD-nH (X^2^ = 0.109, p-value = 0.742). For the On-state session, participants were asked to take their regular medication as prescribed by the neurologist. For the Off DRT state session, participants were asked to take their last dose of dopaminergic medication the evening before testing, no later than midnight, and skip the morning dose, allowing a sufficient time for the washout of the medication (Saranza & Lang, 2021). Long-acting dopaminergic medications (i.e., extended-release formulations) were not taken on the evening before testing. Every testing session started at 9AM. In both sessions, participants started with a resting-state fMRI, followed by the behavioral robotic experiment and the clinical evaluations. The MRI and experimental sessions were conducted approximately 12h after the medications, to minimize lasting medication effects. This approach follows a standardized clinical approach. The investigators were not blinded to the experimental conditions nor the medication status during experiments and the analyses.

### General experimental procedure - riPH in patients with PD

To investigate riPH, we adapted the experimental method and device (Bernasconi et al., 2022) proposed and validated in previous research (Bernasconi et al., 2021). Briefly, sensorimotor stimulation was delivered using with a robotic system consisting of two robotic components (front and back robots) capable of inducing PH. For each experimental session, we applied the following conditions: i) synchronous sensorimotor stimulation, or ii) asynchronous sensorimotor stimulation. In the synchronous condition, patients performed poking movements on the front robot, resulting in sensory feedback on their back via the back robot, that was temporally aligned with the movement. In the asynchronous condition a temporal delay was introduced between the poking movement of the front robot and the sensory feedback via the back robot. During the sensorimotor stimulation, participants were always asked to keep their eyes closed and were exposed to continuous pink noise through headphones, to facilitate immersion in the experience.

### riPH in patients with PD (sensorimotor delay dependency)

After sensorimotor stimulation, participants were asked to report whether they experienced PH or not (Yes/No task), on a trial-by-trial basis. On each sensorimotor stimulation trial, the delay between the movement and the stroking on the back was randomly chosen from a delay of 0 seconds (synchronous condition), 0.250 seconds and 0.500 seconds (asynchronous condition). The trial started with an acoustic signal (400-Hz tone and 0.100 second duration) indicating the beginning of the trial. At this point, the participant started with the poking movements. Once the number of pokes reached a total of ten (automatically counted), two consecutive tones (400 Hz and 0.100 second duration) indicated to the participant to stop the movements and to verbally answer Yes/No to the PH question [question: “Did you feel as if someone was standing close by (behind you or on one side)?”]. The investigators were always placed >4 m away and in front of or on the side of the participants during the experiment. In each session, participants were asked to perform one session of 12 trials (four repetitions per delay).

### Statistical analyses

#### Clinical–demographic variables

Statistical difference between PD-MH and PD-nMH on the measured clinical and demographic variables (cf. Table 1) was assessed using the Welch test, Fisher Exact test, or Mann–Whitney U test, based on normality of the data. Normality was assessed with the Shapiro–Wilk test.

#### riPH in patients with PD (sensorimotor delay dependency)

To investigate how the degree of sensorimotor conflict modulates riPH (Response Yes/No; dependent variable), as a function of Medication (On vs. Off) and Groups (PD-H vs. PD-nH), and Delay (0, 250 and 500 ms), we performed a linear mixed model with a random intercept for each participant. Analyses were performed using the afex package (Singmann et al., 2024) for R. For the analyses, a contrast coding approach was used for factorial variables, and delay was centered (i.e., delays 0, 0.25 and 0.5s were converted to -0.25, 0, and 0.25, respectively). E with the same design were fitted using the ‘brms’ R package (Bürkner, 2017). For these Bayesian regressions, we ran 4 chains of 40000 iterations (including 2000 warmup samples), and we ensured an R-hat close to 1. Priors were set by default according to the brms package in R.

### Neuroimaging analyses

#### Image acquisition parameters

Structural and functional images were acquired at the MRI facility of the Campus Biotech (Geneva, Switzerland), and at the Insel Spital Bern with Siemens MAGNETON Prisma 3T scanners and using a 64-channel head-and-neck coil. An EPI-BOLD sequence was used to acquire functional data (slice thickness=2 mm, FoV=208mm, voxel size=2mm isotropic; TR=700ms; TE=30ms; flip angle=52°; multiband acceleration factor=8; time points=674, approximate acquisition time=8 minutes). Whole-brain T1-weighted anatomical images were acquired using a 3D MPRAGE sequence (slice thickness=1 mm, FoV=256mm; voxel size=1×1×1 mm; TR=2200ms, TE=2.98ms, flip angle=9°, GRAPPA acceleration factor=2).

#### Image processing

Imaging data were preprocessed using CONN (Whitfield-Gabrieli & Nieto-Castanon, 2012) (RRID:SCR_009550) release 22.a (Nieto-Castanon & Whitfield-Gabrieli, 2022) and SPM (Friston, 2007) (RRID:SCR_007037) release 12.7771. Functional and anatomical data were preprocessed using a flexible preprocessing pipeline (default_mnifield, Nieto-Castanon 2020) including creation of voxel-displacement maps, realignment with susceptibility distortion correction using fieldmaps, slice timing correction, outlier detection, indirect segmentation and MNI-space normalization, and smoothing. Functional data were realigned using SPM realign & unwarp procedure (Andersson et al., 2001) integrating fieldmaps for susceptibility distortion correction, where all scans were coregistered to a reference image (first scan of the first session) using a least squares approach and a 6 parameter (rigid body) transformation, and resampled using b-spline interpolation (Friston et al., 1995) to simultaneously correct for motion, magnetic susceptibility geometric distortions, and their interaction. Temporal misalignment between different slices of the functional data was corrected following SPM slice-timing correction (STC) procedure (Henson et al., 1999; Sladky et al., 2011), using sinc temporal interpolation to resample each slice BOLD timeseries to a common mid-acquisition time. Potential outlier scans were identified using ART (Whitfield-Gabrieli et al., 2011) as acquisitions with framewise displacement above 0.9 mm or global BOLD signal changes above 5 standard deviations (Nieto-Castanon, 2022; Power et al., 2014). Functional and anatomical data were coregistered and normalized into standard MNI space, segmented into grey matter, white matter, and CSF tissue classes, and resampled to 2 mm isotropic voxels following an indirect normalization procedure (Calhoun et al., 2017; Nieto-Castanon, 2022) using SPM unified segmentation and normalization algorithm (Ashburner, 2007; Ashburner & Friston, 2005) with the default IXI-549 tissue probability map template. Last, functional data were smoothed using spatial convolution with a Gaussian kernel of 6 mm full width half maximum (FWHM).

In addition, functional data were denoised using a standard denoising pipeline (Nieto-Castanon, 2020a) including the regression of potential confounding effects characterized by white matter timeseries (16 CompCor noise components), CSF timeseries (16 CompCor noise components), motion parameters and their first order derivatives (12 factors) (Friston et al., 1996), outlier scans (below 144 factors) (Power et al., 2014), and linear trends (2 factors) within each functional run, and simultaneous bandpass frequency filtering of the BOLD timeseries (Hallquist et al., 2013) between 0.008 Hz and 0.9 Hz. CompCor (Behzadi et al., 2007; Chai et al., 2012) noise components within white matter and CSF were estimated by computing the average BOLD signal as well as the largest principal components orthogonal to the BOLD average, motion parameters, and outlier scans within each subject’s eroded segmentation masks. From the number of noise terms included in this denoising strategy, the effective degrees of freedom of the BOLD signal after denoising were estimated to range from 1042.9 to 1240.9 (average 1215) across all subjects (Nieto-Castanon, 2022).

#### ROI-to-ROI Functional Connectivity Analysis

ROI-to-ROI connectivity matrices were estimated separately for 1) a network comprising 10 ROIs from PH network and the striatum and 2) the whole-brain functional connectome using 116 ROIs, comprising the Schaefer 100 parcellation (Schaefer et al., 2018) and 16 subcortical ROIs (details below). All ROIs were transposed to the same space than the patients’ functional images. Functional connectivity strength was represented by Fisher-transformed bivariate correlation coefficients from a general linear model (weighted-GLM(Nieto-Castanon, 2020c)), estimated separately for each pair of ROIs, characterizing the association between their BOLD signal timeseries.

PH network and striatum: this consisted of 10 ROIs, including 6 PH network ROIs and 4 striatum ROIs. PH network ROIs were derived from (Bernasconi et al., 2021) and included the left and right inferior frontal gyrus (IFG), left and right posterior middle temporal gyrus (pMTG), and left and right ventral premotor cortex (vPMC). Striatum ROIs were anatomically based on the AAL atlas (Tzourio-Mazoyer et al., 2002) and included left and right putamen (Pu) and left and right caudate (Cd).

Whole-brain functional connectome: this consisted of 116 ROIs, including 100 from Schaefer 100 parcellation (Schaefer et al., 2018), plus 16 additional ROIs including subcortical (left and right thalamus, hippocampus, globus pallidum, amygdala and pons) and striatal regions (left and right putamen, caudate and nucleus accumbens) taken from the Tian subcortical atlas (Tian et al., 2020).

#### Partial Least-Squares Correlation analyses

We used the myPLS toolbox – PLS analyses for medical image processing (https://github.com/danizoeller/myPLS) to perform a Partial Least Squares (PLS) analysis (Krishnan et al., 2011), designed to examine the relationship between brain imaging features and behavioural variables across ON vs. OFF medication status. PLS analyses were performed using the ‘*contrastBehavInteract’* setting of the myPLS toolbox, allowing us to model group contrasts (medication status), behavioural measures (hallucinations in daily life, robot sensitivity), and their interaction.

We conducted two separate partial least squares correlation (PLS) analyses to identify patterns of maximal covariance between brain data and behavioural measures. The first analysis used functional connectivity between a-priori defined ROIs within the PH network and striatum, while the second used whole-brain functional connectivity between ROIs of the Schaefer 116 parcellation. In both cases, brain data were associated with three behavioural variables: medication status (ON vs. OFF), presence of hallucinations in daily life (PD-nH vs. PD-H). To quantify individual performance on the robotic task, we extracted the intercepts from a generalized linear model (GLM) with a quasibinomial distribution, fitted separately for each subject and experimental condition. We prioritized the intercept analysis because our primary behavioral model revealed a significant main effect of medication (see Sensorimotor Delay Dependency section). We used the intercept because it reflects the subject-specific sensitivity irrespective of sensorimotor delay, which was found to be modulated by medication status in the behavioural analyses. Categorical variables (medication status and presence of hallucinations in daily life) were centred around 0 (-0.5 – 0.5). Both behavioural and imaging data were z scored across all subjects. As there were no significant differences in age, disease duration, and sex between PD-H and PH-nH patients (cf. Table 1), these demographic factors were not included in the model, nor regressed out from the brain data.

Brain data (X) consisted of subject-wise brain features derived either from the PH network and striatum or from the Schaefer-116 ROI parcellation. These features formed matrices of dimensions N × M, where N is the number of subjects and M the number of imaging variables - 45 functional connections for the 10 ROIs comprising the PH network and striatum, or 6670 connections for the 116 ROIs of the Schaefer parcellation. Behavioral data (Y) included the three variables described above.

The first step of PLS correlation is the computation of the cross-covariance matrix (R) between X and Y, followed by the singular value decomposition (SVD) of the covariance matrix R.

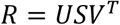

The SVD produces up to three latent variables (LVs), corresponding to the number of behavioral variables, each capturing a distinct pattern of association between brain and behavioral data. Each LV is associated with: (1) a singular value (S) indicating the strength of the brain–behavior correlation explained by that LV; (2) a vector of behavioral loadings (from the corresponding column of U), indicating the relative contribution of each behavioral variable (hallucinations, robot-sensitivity, medication status) and their interaction effects; and (3) a vector of brain loadings (from the corresponding column of V), indicating the contribution of each brain variable (functional connectivity features). Projecting each subject’s original brain data (X) onto the brain loadings (V) yielded subject-wise “brain scores”, reflecting the similarity of each subject’s brain pattern to the latent brain pattern. Likewise, projecting behavioral data (Y) onto the behavioral saliences (U) produced “design scores”, reflecting how closely each subject’s behavioral profile matched the latent behavioral pattern. The correlation between brain scores and design scores was used to assess the coherence and validity of the significant LV, and the patterns of association across medication status and presence of hallucinations in daily life

To determine whether each LV explained significant brain–behavior covariance, we applied permutation testing with 10,000 permutations to generate a null distribution of singular values. A LV was considered significant if its singular value exceeded the 95th percentile of this null distribution. For significant LVs, the robustness of brain and behavioral loadings was evaluated using a bootstrap procedure with 5,000 resamples with replacement. Loadings were recalculated for each bootstrap sample, providing bootstrap distributions for estimating the stability of the brain–behavior relationships. Behavioural and imaging loadings were considered statistically significant when the 95% confidence intervals derived from the bootstrap samples did not include zero, indicating stable contributions across bootstrap samples.

#### Network-pair enrichment analysis

For the whole-brain PLS analysis, we summarized the distribution of significant imaging loadings across the 7 canonical resting-state networks (RSNs;(Yeo et al., 2011)), subcortical regions, and striatum. For each network pair (within-and between-network), we computed the percentage of significant edges with positive loadings. Because raw percentages are disproportionately unstable for networks with few nodes (e.g., the Limbic network), we assessed network-pair enrichment relative to chance using a permutation-based approach: network labels were randomly shuffled 5,000 times while preserving network sizes, and the percentage of significant positive edges was recalculated for each network pair under each permutation, generating a null distribution. For each network pair, a one-tailed z-score was computed as the deviation of the observed percentage from the mean of its corresponding null distribution, expressed in units of the null distribution’s standard deviation. Network pairs were considered significantly enriched when z > 1.645 (p<.05, one-tailed). Given the low absolute number of negative loadings across the whole-brain analysis (25/327 significant edges, 7.6%), the same enrichment procedure was not applied to negative loadings; raw percentages and absolute edge counts per network pair are instead reported descriptively for both positive and negative loadings in Supplementary Figure S1.

## Data Availability

All data produced in the present study are available upon reasonable request to the authors.

## Acknowledgments

The authors disclosed receipt of the following financial support for the research, authorship, and/or publication of this article: This project was supported by two generous donors advised by CARIGEST SA, the first one wishing to remain anonymous and second one being Fondazione Teofilo Rossi di Montelera e di Premuda to O.B.; Catalyst the Bertarelli Foundation to O.B; Empiris foundation to O.B; Parkinson Schweiz to O.B and F.B.; Synapsis Foundation to O.B and F.B.; Leenaards foundation to F.B, Swiss National Science foundation (n° 320030_188798), EPFL Neuro X Post-doctoral fellowship program to S.S.

## Authors contribution

Conceptualization: F.B.,S.S., O.B.; O.B & F.B designed the experiment.

Formal analysis: F.B.,S.S., L.B

Investigation: F.B., J.P, M.M, S.H.A., S.C.

Resources: S.S., J.P., F.B., O.B.

Data curation: F.B., S.S., L.B., J.P.

Writing – Original Draft: F.B., S.S., O.B.

Writing – Review & Editing: all the co-authors.

Visualization: F.B., S.S.

Supervision: F.B., S.S., O.B.

Project administration: F.B., S.S., O.B.

Funding acquisition: F.B., S.S., O.B.

