## Supplementary materials for "Dopaminergic therapy selectively amplifies hallucination susceptibility in patients with Parkinson’s disease with cortico-striatal hyperconnectivity"

*Demographical, clinical, and neuropsychological data*

In the PD-H subgroup, 7 patients reported passage hallucinations, 5 presence hallucinations, 6 visual illusions/pareidolia, 2 complex visual hallucinations, 5 auditory and 3 tactile, consistent with the habitual prevalence of the different hallucinations in PD (Bernasconi et al., 2021, 2023, Albert et al., 2024). A single patient can experience more than one hallucination.

*Sensorimotor delays and dopamine induced modulations of PH-like hallucinations in PD patients with hallucinatory trait*

| Effect | Chisq | p-value |
| --- | --- | --- |
| Delay | 27.915 | < 0.001 |
| Group | 2.383 | 0.123 |
| Medication | 2.075 | 0.150 |
| Delay:Group | 0.008 | 0.927 |
| Delay:Medication | 0.887 | 0.346 |
| Group:Medication | 1.514 | 0.219 |
| Delay:Group:Medication | 4.044 | 0.044 |

**Table S1.** Results of the linear mixed model assessing statistical differences in response (dependent variable), with delay, medication, and groups as fixed effects. An interaction between the three terms was included, as well as a random intercept per subject.

| Effect | Chisq | p-value |
| --- | --- | --- |
| Delay | 24.160 | 0.000 |
| Medication | 6.984 | 0.008 |
| Delay:Medication | 1.026 | 0.311 |

**Table S2.** Post-hoc results of the linear mixed model for PD-H assessing statistical differences in response (dependent variable), with delay and medication as fixed effects. An interaction between the three terms was included, as well as a random intercept per subject.

| Effect | Chisq | p-value |
| --- | --- | --- |
| Delay | 9.344 | 0.002 |
| Medication | 0.011 | 0.918 |
| Delay:Medication | 3.008 | 0.083 |

**Table S3.** Post-hoc results of the linear mixed model for PD-nH assessing statistical differences in response (dependent variable), with delay and medication as fixed effects. An interaction between the three terms was included, as well as a random intercept per subject.

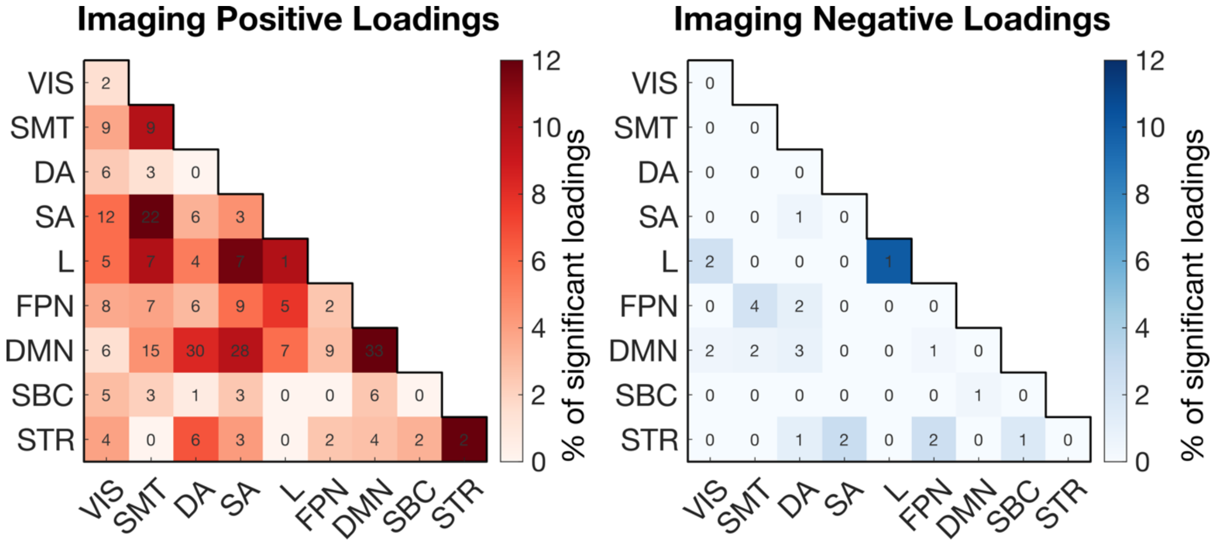


**Supplementary Figure S1. Descriptive network-pair distribution of positive and negative significant imaging loadings (whole-brain PLS analysis).** Matrices show the percentage of significant imaging loadings across network pairs for the whole-brain latent component (LC) reported in Figure 4, separately for positive (red) and negative (blue) loadings. For each network pair, values indicate the percentage of connections with statistically reliable loadings across bootstrap samples (95% confidence interval not including zero), out of the total possible connections within that network pair; numbers within each cell indicate the corresponding absolute edge count. Matrices are organized by the 7 canonical resting-state networks (RSNs), subcortical regions, and striatum: VIS = Visual Network; SMT = Somatomotor Network; DA = Dorsal Attention Network; SA = Salience Network; L = Limbic Network; FPN = Fronto-Parietal Network; DMN = Default Mode Network; SBC = Subcortical regions; STR = Striatum (including left and right putamen and caudate). Unlike the network-pair enrichment analysis reported in Figure 4B, these percentages are shown descriptively, without permutation-based statistical testing. This is because the total number of negative loadings was low (25/327 significant edges, 7.6% of all significant edges), resulting in most network-pair blocks containing very few negative edges (median = 0–1 edges per block), which precludes reliable enrichment testing at the network-pair level for the negative loadings panel. The corresponding positive loadings panel is shown here for direct visual comparison with the negative loadings panel and reproduces the same raw percentages underlying the statistically-tested enrichment matrix in Figure 4B.
